# Cannabinoids For Fibromyalgia: An Updated Systematic Review

**DOI:** 10.1101/2022.05.17.22275200

**Authors:** Jean Claude Scicluna, Giuseppe Di Giovanni

**Author notes:** Corresponding authors: Jean Claude Scicluna, Giuseppe Di Giovanni.

## Abstract

Fibromyalgia is an increasingly prevalent condition resulting in high morbidity and economic burden for sufferers. Minimal to modest benefit has been achieved by pharmacotherapies, creating a strong rationale for novel therapies. Substantial evidence has implicated the endocannabinoid system in the modulation of fibromyalgia symptoms. However, the therapeutic potential and potential adverse effects of cannabis-based therapy in fibromyalgia are still under-reported, leading to clinicians’ hesitation to opt for such therapy. This systematic review examined the literature and provided a critical review of the safety and efficacy of cannabis-based therapy in fibromyalgia. It resulted that medical cannabis is a safe and effective treatment option for fibromyalgia, whilst further research in this area is needed.

## INTRODUCTION

Fibromyalgia is a relatively common disorder whose primary manifestation is chronic widespread pain(Bair & Krebs, 2020). Prevalence has increased dramatically with the development of more sensitive diagnostic criteria throughout the years, with a strong preponderance being seen in women, although with the most recent modified the American College of Rheumatology (ACR) criteria, numbers in men are also strong on the increase (Jones et al., 2015).

Fibromyalgia carries a significant economic burden, not only because of the functional impairments to the patient’s work and domestic life but also due to the cost of health services. Indeed, the yearly cost of these health services is for example about a thousand euros as revealed by a Dutch study (Boonen et al., 2005 Prior pain conditions are strongly associated with fibromyalgia, possibly due to activation of secondary central sensitization (Clauw, 2015). Indeed, patients with fibromyalgia tend to also suffer from conditions such as chronic low back pain, irritable bowel syndrome, temporomandibular disorders, as well as sleep disorders, anxiety and depression

The overall approach to treating fibromyalgia is focused on maintaining or improving function, improving quality of life, and managing symptoms(Bair & Krebs, 2020; Løge-Hagen et al., 2019) et al., 2019). The mainstay of treatment has so far been focused on non-pharmacologic areas such as patient education, maintenance of sleep hygiene, a balanced diet, regular physical activity and an overall healthy lifestyle. Psychological treatments such as cognitive behavior therapy (CBT) have shown modest results in improving mood, alleviating pain and improving disability(Bernardy et al., 2013). Moreover, pharmacologic therapies have so far shown minimal to modest benefit in fibromyalgia patients, with simple analgesics such as acetaminophen and non-steroidal not being found effective,(Derry et al., 2017) and whilst gabapentinoids and tramadol have been used by clinicians to some reported effect, although a Cochrane review of studies concludes that there is inconclusive evidence for gabapentin reducing pain in fibromyalgia (Cooper et al., 2017). Opioids other than tramadol (due to its serotonergic and noradrenergic uptake properties) have also not been found effective in fibromyalgia, despite clinicians’ ongoing use of these drugs in practice (Goldenberg et al., 2016). Duloxetine, a selective serotonin and noradrenaline reuptake inhibitor (SSNRI) antidepressant, has been shown to improve symptom scores for pain and fatigue (but not insomnia) compared to placebo, however, adverse effects resulted in high dropout rates (Häuser et al., 2013.)

In light of all these efforts, there is a strong rationale surface for the investigation of novel pharmacologic therapies which can be added to the clinicians’ arsenal.

The use of the plant *cannabis sativa* has been linked with medicinal and recreational purposes for millennia. Since the discovery and characterization of the endogenous cannabinoid system, several studies have evaluated how cannabinoid compounds and, particularly, how the modulation of the endocannabinoid (eCB) system influences a wide range of functions, from metabolic to mental disorders (Bahji et al., 2020; Blessing et al., 2015; Bonaccorso et al., 2019; Breuer et al., 2016; Campos et al., 2013; Crippa et al., 2011; Faraji et al., n.d.; Hill et al., 2009; Linares et al., 2019; Lisboa et al., 2017; Rosenberg et al., 2017; Scarante et al., 2017; Schier et al., 2012; Sethi et al., n.d.; Yohn et al., 2017 (Korem et al., 2016). It was demonstrated in the 1980s that the effects of cannabinoids were mediated by their interaction with specific sites, resulting in the activation of a G protein signaling and the inhibition of adenylate cyclase activity (Howlett, 1987). The eCB neuromodulator system regulates emotional, cognitive, neurovegetative and motivational processes. Crucial to the functioning of the endocannabinoid system are the cannabinoid 1 and 2 receptors (CB1 and CB2Rs), which have been found in high density throughout various areas of the brain, especially the prefrontal cortex and hippocampus). Substantial evidence has accumulated implicating a deficit in eCB in the etiology of depression; accordingly, pharmacological augmentation of endocannabinoid signaling could be a novel target for the pharmacotherapy of depression (Hill et al., 2009)(

Whilst some studies suggest a role for endocannabinoid deficiency in pain and specifically, in fibromyalgia the role of endogenous cannabinoids remains unclear and more studies are required in this regard. This is due to the complexity of both the eCB system and pain, a “multidimensional, dynamic integration among physiological, psychological, and social factors that reciprocally influence one another”. (Meints & Edwards, 2018) In addition, cannabinoids may attenuate low□grade inflammation, another postulate for pathogenesis in people with fibromyalgia (Üçeyler et al., 2011). Cannabinoids may reduce peripheral and central sensitization of nociception by altering cognitive and autonomic processing of chronic pain ((Guindon et al., 2009). The distribution of CBRs in the frontal□limbic seems to point toward cannabinoids preferential targeting the affective qualities of pain, known to have an important contribution to the suffering of patients with fibromyalgia (Lee et al., 2012). Finally, considering that fibromyalgia is a stress□related disorder(van Houdenhove & Egle, 2004), cannabinoids might be helpful in buffering stress and modulating emotional and cognitive functions (Colangeli et al., 2021). Apart from phytocannabinoids, boosting eCBs such as anandamide (AEA) and 2-Arachidonoylglycerol (2-AG) by blocking the specific catabolic enzymes i.e., fatty acid amide hydrolase (FAAH) and monoacylglycerol lipase (MAGL), is being tested as a novel treatment for pain (Papa et al., 202). Therefore, taking into consideration the complexity of symptom expression and the absence of ideal treatment, the potential for manipulation of the cannabinoid system as a therapeutic modality is attractive. Future research into the clinical utility of eCB metabolism manipulation in fibromyalgia is also expected.

With the aim of analyzing the current evidence on the use of cannabinoids in fibromyalgia we, therefore, have run a systematic review of the clinical study.

## SEARCH METHODS AND SELECTION CRITERIA

The PRSIMA (Preferred Reporting Items for Systematic Review and Meta-Analyses) guidelines were followed for writing this review. PubMed/Medline were identified as databases for the research. A search was conducted in March 2022 using the keywords “cannabis + fibromyalgia” and “cannabinoids + fibromyalgia”. The following inclusion criteria were used in the selection of studies: a) existing translation in the English Language b) published from 2008 to 2022 c) Randomized Controlled Trials, observational studies, retrospective studies and comparative studies d) minimum of 15 patients in each study e) any preparation of synthetic cannabinoid or herbal cannabis.

## RESULTS

The search resulted in 31 and 33 results on PubMed for each search term pair respectively, but after removal of duplicates and studies which did not meet study criteria, 8 articles were deemed relevant. One study using fibromyalgia as adjunctive therapy to mainstream therapy was used (see Fig. 1). The references of these studies were also reviewed for additional studies, and 1 further study was found in this way. The aim was to provide an in-depth screen of the current literature surrounding the action of cannabis and cannabinoid therapies in the outcomes of Fibromyalgia patients, whilst only including studies with validated and reliable methods of data collection, and avoiding anecdotal studies. Table 1 shows a summary of the selected studies and the criteria used to provide this analysis.

## DISCUSSION

### Critical Analysis

This review included 7 studies using herbal cannabis preparations (phytocannabinoids) ((Fiz et al., 2011; Giorgi et al, 2019.; Habib & Artul, 2018; Sagy et al., 2019; van de Donk et al., 2019; Yassin et al., 2019))(Chaves et al., 2020a)) and 2 studies using the synthetic cannabinoid, nabilone (Skrabek et al., 2008; Ware et al., 2010)) in patients with fibromyalgia. The phytocannabinoid preparations were a variety of THC-dominant cannabis flowers and oil extracts.

From the outset, it becomes evident that the reliability of studies using herbal cannabis suffered because of the lack of consistency of cannabis used. In all but one of the studies (van de Donk et al., 2019), we were unable to identify the strain, potency, cannabinoid profile and terpene profile within the formulation. Only three studies using herbal cannabis included formulations with cannabidiol (CBD) content(Chaves et al., 2020; Giorgi et al, 2019.; van de Donk et al., 2019)), and only two included CBD in significant amounts (1:1 ratio with THC) (Giorgi et al, 2019.; van de Donk et al., 2019)).

### The dosage varied widely throughout the studies

These problems were not encountered in the two studies using the synthetic delta-9 THC analogue Nabilone(Skrabek et al., 2008; Ware et al., 2010)(Skrabek et al., 2008; Ware et al., 2010)(Skrabek et al., 2008; Ware et al., 2010)(Skrabek et al., 2008; Ware et al., 2010). The re-emergence of plant medicine in the 21^st^ century has so far not been met with sufficient innovations in clinical research attempting to gain quantitative insight into the benefits of this section of medicine. Pharmaceutical markets are now embracing the change of consumer mentality surrounding the ingestion of lab-made synthetic compounds towards more natural products. It is imperative that this shift is not simply a question of marketing aesthetics, but also viable data.

### Baseline demographics/comorbidities

There was a wide variety of patient characteristics throughout all studies. Most studies showed a prevalence of women, with (Chaves et al., 2020) having a patient cohort consisting of only women. The most abundant cohort by far was women aged 30 to 50 years (Chaves et al., 2020b; Fiz et al., 2011; Habib & Artul, 2018; Sagy, Schleider, et al., 2019; Skrabek et al., 2008; v. Giorgi1 et al., n.d.; van de Donk et al., 2019; Yassin et al., 2019))(Ware et al., 2010)(Ware et al., 2010)(Ware et al., 2010)(Ware et al., 2010). Women with fibromyalgia are notably younger than men, and are more likely to be suffer from comorbid psychiatric or connective tissue disorders, whilst men often had multiple medical comorbidities(Sagy, Schleider, et al., 2019). Fibromyalgia shows a gender-dependent, multi-modality of symptoms, and this should be considered in the treatment approach (Arout et al., 2018). Of the studies analyzed in this review, only one study had sufficient recruitment to allow meaningful observations to be made in males(Sagy, Schleider, et al., 2019). The biological mechanisms of interaction of the eCB system differ between sexes, notably how throughout most of the brain, eCB content is estrous cycle-dependent in females in comparison to males, as shown by preclinical studies (Bradshaw et al., 2006). Therefore, the results should not be generalizable between the sexes.

In future studies, findings should be further specified into reference groups according to similar characteristics, both in comorbidities, age and sex demographics.

Post-hoc analysis of the effect of the baseline characteristics on the efficacy of the treatment should be a target for future research endeavors.

### Inclusion/exclusion criteria

There were inconsistencies in the inclusion and exclusion criteria between the studies. Some only used fibromyalgia patients with moderate to severe symptoms (Chaves et al,2020), whilst others excluded patients using any other pain medication than mild analgesics (Van de Donk et al, 2019). One study (Habib & Artul, 2018) excluded patients with malignancy-associated or rheumatic-associated comorbidities.

Exclusion criteria were more generalizable and frequently included the history of psychotic symptoms or psychosis-associated conditions such as schizophrenia and diagnoses which could explain symptoms other than fibromyalgia.

### Route of administration

Route of administration of the cannabinoids varied across studies and included smoking, vaporization, ingestion of oil drops or a combination of these. Dosage amount and strategy were also widely varied. There is some evidence in the literature to show that some methods of administration have benefits and risks with respect to each other (Aston et al,2019),(Russo, 2016b) however there is a lack of studies comparing the benefits and side effect profile of the different routes of administration (ROAs). The general consensus points to vaporization being the safest option, with improved beneficial effects and reduced adverse profiles. This contrasts with smoking, one of the studies (Habib & Artul,2018) showed that participants using smoking as their main ROA had a greater incidence and severity of adverse effects such as the dry mouth and red eyes. Respiratory problems such as coughing, wheezing and increased phlegm production have been proven to be associated with smoking in various reviews (Peiffer et al,2018).

It is pivotal for future studies, as well as the future of herbal cannabis preparations in mainstream medicine, that more investigation into ROAs is done. The aim should be the provision of measurable and reliable dose titration, whilst maximizing therapeutic benefit and without causing pulmonary damage.

### Drug interaction/comparison with other analgesic treatments

The studies selected did not identify any possible drug interactions or their effects in both the short term and long term. Fibromyalgia patients are often highly comorbid, and tend to use many different forms of concurrent analgesia in an attempt to better their symptoms.

Moreover, only one study (Ware et al., 2010) directly compared cannabinoids to mainstream medication options such as amitriptyline, a tricyclic antidepressant, showing the synthetic cannabinoid nabilone to be favorable in both efficacy and side effect profile. It is crucial that in future, studies (including those using phytocannabinoids or herbal cannabis preparations) are compared with other standard treatments for pain management so as to provide reliable comparison data. This allows an informed discussion over the pros and cons of each treatment in the clinical setting, empowering fibromyalgia patients to make informed choices about their medication as well as providing guidance to medical professionals.

### Diagnostic validity

7 of the 9 studies(Chaves et al., 2020; Fiz et al., 2011; Giorgi et al, 2019.; Sagy, Schleider, et al., 2019; Skrabek et al., 2008; van de Donk et al., 2019; Ware et al., 2010)) used established criteria for diagnosis in the recruitment of patients. The criteria used in 5 of the 7 studies were the ACR 2010 criteria.2 studies used the American College of Rheumatology 1990 criteria(Skrabek et al., 2008; Ware et al., 2010)(Skrabek et al., 2008; Ware et al., 2010)(Skrabek et al., 2008; Ware et al., 2010)(Skrabek et al., 2008; Ware et al., 2010) Recommendations for future studies include using recognized criteria for verification of a fibromyalgia diagnosis between physicians. A complicating factor in this context would be that the precise parameters of a fibromyalgia diagnosis are operationalized in an entirely different way between cultures, medical bodies and hospital systems.

### Assessment outcome

The methods used for the assessment of outcomes in patients varied. Chaves et al. (2020) used the Fibromyalgia Impact Questionnaire (FIQ), an instrument validated for use in this condition (Bennett et al., 2009) to investigate outcomes, whilst Van De Donk et al (2019) took a completely different approach by comparing spontaneous pain scores and electrical pain thresholds. Similar to Chaves et al. (2020), Yessin et al (2019), Habib & Artul (2018), Ware et al. (2010), (Fiz et al., 2011), Giorgi et al (2019) and (Skrabek et al., 2008) all used validated instruments for assessing the severity of symptoms in fibromyalgia. Moreover, all but Fiz (2011) used a medium-term approach (weeks to months) for the assessment of outcomes. Fiz (2011) assessed short-term outcomes, as did Van De Donk (2019).

The studies using similar instruments for outcome reportage allowed trends to be observed. Some studies explored other experimental designs, this made a meta-analysis of the data less likely to reach definitive conclusions. It is important for future studies to operationalize the specific type of pain that is being treated for and assessed in fibromyalgia.

It is helpful in the pursuit of reliable clinical data that similar instruments are used to allow cross-referencing and meta-analysis of the results obtained. This allows both studies and reviews to give better peace of mind to clinicians and other professionals using this data to help patients make informed choices about their medication choices.

### Participant education

It was noted that in the selected studies, little to no focus was placed on participant education. Indeed, it was seen from characterization studies allowing more freedom for patients to choose preferential methodologies of cannabis use, a wide variety of methods were employed by patients to use their medicine. In more organised study designs, no reference was made to teaching patients about the use of the delivery devices and signs of intoxication. Whilst it can be argued that this prevented possible interviewer bias, it is reasonable to assume this had a bearing on results. Patient education is a vital factor in the medical and pharmaceutical industries and will be even more so in the future as the culture of shared decision-making and patient-centric care takes precedence in clinical settings. Patient education should not be limited to naïve users, as even experienced cannabis users tend to derive most of their knowledge from their own experiences, and this knowledge tends to show discrepancies from available evidence (Kruger et al, 2020).

It is notable that several participants who dropped out from the initial phases of some studies (Ware et al,2010) (Skrabek,2008) mentioned the stigma surrounding cannabis as a factor. The data from more recent studies do not explore whether stigma continues to be a factor affecting treatment outcomes. This is an important consideration for future studies, especially those exploring pain syndromes and cannabis.

### Participant blinding

In studies that incorporated blinding of participants, challenges were generally faced by those using THC-dominant herbal cannabis preparations. Indeed, Van De Donk et al (2019) used Bang’s Blinding index (Bang et al,2004) to show that Bedrocan faced the biggest issues with respect to blinding, however, successful blinding was still possible according to the blinding index’s cut off rate (<0.5). The lack of psychoactive effects with placebo compared to potent THC-dominant strains is a further challenge for clinical trials, possibly leading to an overestimation of the THC-dominant preparations’ beneficial and/or adverse effects.

These issues are less problematic in the study of non-psychoactive cannabinoids, which constitute a large majority of the phytocannabinoids found in nature.

### Adverse effects (baseline, assessment of severity)

In all 9 studies included in this review (Chaves et al., 2020b; Fiz et al., 2011; Habib & Artul, 2018; Sagy, Schleider, et al., 2019; Skrabek et al., 2008; v. Giorgi1 et al., n.d.; van de Donk et al., 2019; Ware et al., 2010; Yassin et al., 2019), adverse effects were mild to moderate and the major adverse effects seen were drowsiness and a dry mouth. Other adverse effects reported in a minority of the participants included mild dizziness, red eyes, hunger and palpitations. Studies using smoking as a route of administration also included sore throat and coughing as immediate side effects. This was also seen in other inhalation methods such as vaporization. Nausea was generally observed in ingestible oil preparations.

Van De Donk et al (2019) was the only study where patients reported a significant drug high, whilst the experience of the psychoactive effects in patients was not explored in detail. This was only seen with a THC dominant strain Bedrocan, and not with CBD dominant strain Bedrolite or Bediol, which contains CBD:THC in a ratio of approximately 1:1. Patients recruited for this study were cannabinoid naïve and instructed to fast before the study. It is unclear if this is significant to the adverse effect findings. Together with the in-depth, jargon-free participant education described above, more research into the psychoactive actions of THC and the milieu of interacting factors is necessary for THC to be incorporated safely into medical use.

Furthermore, future studies should aim to create a baseline for symptoms commonly seen in fibromyalgia, so as to distinguish them from adverse reactions due to medical cannabis use. For example, symptoms such as dizziness, nausea, constipation, dry eyes and dry mouth are consistent in both (Van De Donk et al (2019). Additionally, whilst some studies did include a control group, future studies should control for cannabis use patterns amongst patients, such as naïve patients, past users, current users, and non-users. Ideally, future studies should control for medical and recreational use since the participant backgrounds and motives surrounding cannabis use in these two groups might vary and cause significant interference in the results.

## CONCLUSIONS & FINAL RECOMMENDATIONS

In conclusion, the current evidence is indicating that cannabinoids have the potential to be a safe and effective treatment modality for patients with fibromyalgia. The significant limitations of the current body of evidence prevent more definitive statements and more widespread application of cannabinoids in a clinical setting. The shining potential of this treatment option to provide much-needed, safe effective relief in these patients, together with these limitations, provide a strong rationale for further studies into the subject.

In summary, recommendations for future studies include stratification of findings into reference groups according to similar characteristics, as well as a post-hoc analysis of the effect of the baseline characteristics on the treatment outcomes. Future research should include an analysis of the effect of ROAs, more insight into common and uncommon drug interactions of cannabinoids and the effect of concurrent analgesia. There is a need for more generalizability across studies, and instruments used for outcome reportage should be standardized. Inclusion/exclusion criteria also require more standardization, whilst ensuring that no form or severity of fibromyalgia is excluded, and that established criteria are used for widespread diagnostic validity. Adverse effect profiles need further analysis, with larger sample sizes and classified according to a baseline of the present symptoms of fibromyalgia. More work needs to be done with respect to the challenges of effective participant blinding when using cannabinoids in clinical research.

The legal climate surrounding medical and recreational cannabis use is dynamic and changing rapidly, especially in the European Union where we are seeing a swathe of member states regulating the use of cannabinoids in medical, but also cosmetic, recreational, and nutritional contexts. Policymakers would be wise to identify clinical research as a pivotal starting point for these measures and make decisions based on the best evidence. It is vital also for regulators and governments to ensure market concerns do not out-pace research. Finally, patient education and patient attitudes towards cannabinoids, their benefits and risks, should be a backbone of future clinical integration of these substances. Innovations in medical science are moot if not translated, accurately and simply, into a widespread culture of responsible use of medicines in our societies.

## Supporting information

Fig.1 Study Flow Diagram

Fig. 2 Overview of results

## Data Availability

All data produced in the present work are contained in the manuscript.

## ACKNOWLEDGEMENTS

We thank Malta Enterprise for supporting our research on cannabinoids in fibromyalgia.

