## Supplementary figures and images for "Cannabinoids For Fibromyalgia: An Updated Systematic Review"

### Fig.1 Study Flow Diagram

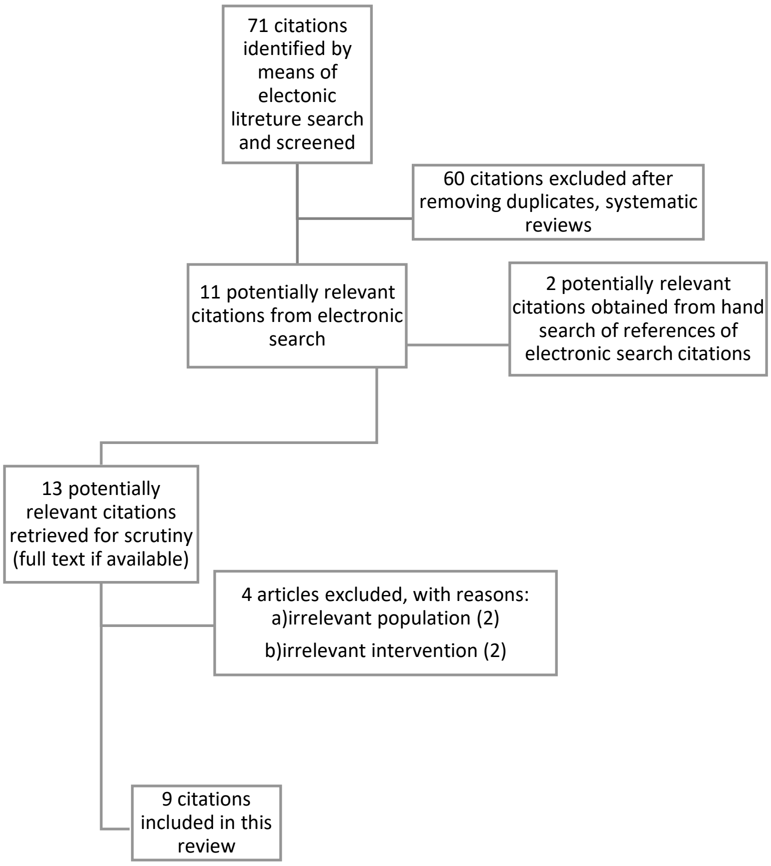


**Figure 1**

Study flow diagram.
