## Supplementary material for "Cannabinoids For Fibromyalgia: An Updated Systematic Review": Fig. 2 Overview of results

| Main Author (s) | **Type of study** | | **Number of participants** | | **Duration of Assessment** | | **Type of Cannabinoid agent used** | | **Dose** | **ROA** | **Adverse events reported** | **Treatment Outcomes** |
| --- | --- | --- | --- | --- | --- | --- | --- | --- | --- | --- | --- | --- |
| Skrabek et al (2008) | Randomized, Double-Blind Placebo Controlled Clinical Trial | | 40 | | 4 weeks | | Nabilone, Synthetic cannabinoid | | 0.5mgOD- 1mg BD | Ingestion | Drowsiness, dry mouth |  |
| Ware et al (2010) | Randomized, Double Blind,active-control, equivalency crossover clinical trial | | 31 | | 2 weeks | | Nabilone, Synthetic cannabinoid | | 0.5mg OD 0 1mg OD | Ingestion | Drowsiness | Pain alleviation, improved QoL, improved mood comparable to Amitriptyline. Improved sleep superiorly to Amitriptyline |
| Fiz et al (2011) | Observational Cross sectional Study | | 28 | | 2 hours | | THC dominant flower unspecified strain, unofficial sources | | Varied, unreliable | Inhalation (smoking), ingestion, combined | Drowsiness, dry mouth | Strong relief for pain, sleep disturbance, stiffness, mood disorders and anxiety. Mild relief for headaches |
| Habib & Artul (2018) | Retrospective Online Self Report Survery | | 26 | | N/A | | THC dominant flower and oil extract, unspecified strain | | 17.7-34.6g per month | Inhalation (smoking) 58% vaporisation (23%), conbined (14%), ingestion (8%) | Dry mouth (27%), red eyes(27%) and hunger (15%) | Increased capacity for work in 46% of participants |
| Van de Donk et al (2019) | Randomized, placebo Controlled 4-way crossover clinical trial | | 20 | | 3 hours | | Bedrocan, Bedrolite, Bediol medical cannabis flowers, Placebo cannabis | | 22.4mg Bedrocan, 18.4mg, Bedrolite, 13.4mg Bediol STAT | Inhalation (vaporisation) 100% | Drug high (80%) for Bedrocan, coughing (70%),nausea and dizziness (15%) and sore throat (10%) | THC correlate with increased pain threshold. Bediol resulted in 30% in pain scores |
| Sagy et al (2019) | Prospective, Observational Study | | 367 | | 6 months | | THC dominant flower, unspecified strain | | 670mg-1000mg daily | Inhalation (smoking) & ingestion | Mild dizziness 7.9%), Dry mouth (6.7%) GI symptoms (5.4%) | Reducedpain (44%), Better sleep (73.4%), Depression scores improves (80.8%) |
| Yassin et al (2019) | Observational, Cross-Over Study | | 31 | | 3-6months | | THC dominant flower, unspecified strain | | 20g monthly | Inhalation (smoking), Inhalation (vaporisation) | Red eyes (90%), Constipation (50%) | Decreased pain intensity, Increased ROM |
| Giorgi et al, (2019) | Prospective Observational Study | 102 | | 6 months | | Bedrocan and Bediol | |  | | Drops | Dizziness (21%), sleepiness (16%), palpitations (12%), nausea (9%) and dry mouth(9%). | 30% improvement in PSQI and FIQR scores |
| Chaves et al (2020) | Randomized, Double-Blind, Placebo- Controlled Clinical Trial | 17 | | 8 weeks | | THC dominant /trace CBD oil | | 1.2mg THC/ 0.02mg CBD to 4.4mg THC/0.08mg CBD daily | | Ingestion | Somnolence | Improved QoL,pain reduction, improved functionality, improved sense of well being |

Table 1 shows``` a summary of the selected studies and the criteria used to provide this analysis.
